# Stakeholder perceptions of the VIrtual Physiotherapist-led Evaluation of low back pain Referrals to spine surgeons (VIPER) model of care: a qualitative study

**DOI:** 10.1101/2025.10.29.25338508

**Authors:** Tarcisio F de Campos, Andrew R Gamble, Giovanni E Ferreira, Christopher G Maher, David B Anderson, Joshua M Hutton, Sophie MacPherson, Christopher S Han, Mouna Sawan, Ian Harris, Sam Adie, Leanne Hassett, Christopher Legg, James Van Gelder, Mark Halliday, Monika Boogs, Rowena Charteris, Christopher Williams, Laurent Billot, Deborah Fullwood, Edward Riley-Gibson, Emma Molineux, Karen Tambree, Maria Tchan, Saurab Sharma, Abby Haynes, Jemma Moujalli, Edith Wong, Joshua R Zadro

## Abstract

**Introduction:** Despite guidelines advising that non-serious low back pain (LBP) should be managed with self-management advice and exercise, referrals to spine surgeons are common. High referral rates to public hospital spine surgery clinics means many surgeons can’t assess new cases within 1-2 years. In many cases, patients referred to these clinics have not tried recommended non-surgical care, and while they wait for surgical review, they develop symptoms which are complex and costly to manage. We developed the VIrtual Physiotherapist-led Evaluation of low back pain Referrals to spine surgeons (VIPER) model of care to help clinics identify new referrals who can be managed sooner by a physiotherapist and reduce wait times for those needing surgical review.

**Aim:** To qualitatively explore stakeholder perceptions of the VIPER model of care, as part of a broader program of work to co-design VIPER and then evaluate it in a large, multi-site randomised controlled trial.

**Methods:** We conducted semi-structured interviews with people with LBP (including those with lived experience of being referred to spine surgery clinics), clinicians who manage patients with LBP (e.g., physiotherapists, spine surgeons), and other key stakeholders (e.g., physiotherapy and spine surgery departments managers). Participants were recruited via social media advertisements, the authors’ networks, and snowball sampling. Participants completed pre-interview questionnaires capturing data to support purposive sampling based on demographics, symptoms and professional characteristics. Interview transcripts were analysed using an inductive descriptive qualitative analysis.

**Results:** Interviews with 39 participants (6 people with LBP, 26 clinicians, and 7 other key stakeholders) highlighted four key themes: 1) current gaps in LBP care pathways and implementation considerations (covering need for appropriate patient and clinician education, referral inefficiencies, coordination challenges, and access barriers); 2) perceptions of the role of physiotherapy in LBP care and patient selection for VIPER; 3) support for VIPER as a means to improve patient outcomes and health system efficiency; and 4) views on virtual assessment and escalation, recognising the value of hybrid models and its limitations.

**Conclusion:** The proposed VIPER model of care appears feasible, acceptable, and well-suited to improve LBP care by promoting guideline-based non-surgical management and reducing wait times for surgical review among those who need it most. The virtual component of the model offers flexible, patient-centred delivery with potential system-wide benefits, supporting further piloting and evaluation, and possibly wider applications in musculoskeletal care.

## INTRODUCTION

Low back pain (LBP) is the leading cause of disability in Australia and globally, and affects over 4 million Australians.^1^ Clinical practice guidelines recommend that most people with non-serious low back pain (e.g., non-specific LBP, mild and stable radiculopathy; >90% of back pain in primary care) should be managed with advice and education to support self-management and exercise therapy.^2^ Guidelines also advise against surgery for non-serious LBP given evidence that surgery is not superior to non-surgical care and may cause harm.^2^ Despite this, referrals to spine surgeons are common; each year 45,000 lumbar spine surgeries are performed in Australia,^3^ costing $1 billion,^3^ and at least 115,000 people referred to spine surgeons are deemed not suitable for surgery.^4–6^

High referral rates to spine surgery clinics, particularly in the public system, mean many surgeons can’t assess new non-serious cases within 1-2 years;^7^ ^8^ ^9^ these cases make up ∼50% of referrals.^7^ ^9^ In many cases these patients have not tried recommended non-surgical care, and while they wait 1-2 years to see a surgeon, they develop complex symptoms which are costly to treat.^10^ This problem is often worse in rural, remote and regional areas where waiting times are even longer due to large catchment areas and a low number of spine surgery clinics.^11^

A promising solution to this health service issue is a triage and management model of care where physiotherapists intercept new referrals for non-serious LBP to spine surgery clinics and determine their suitability for surgical review or a course of non-surgical care. Similar models have been explored in Australia. For example, an observational study in a Victorian Hospital found 2 in 3 new orthopaedic referrals for musculoskeletal conditions could be effectively managed by a physiotherapist without needing to see a surgeon.^12^ However, systematic reviews^13^ show that there has been no high-quality clinical trial evaluating the effectiveness and cost-effectiveness of this model of care and no one has explored a virtual service to ensure equitable care access outside of metropolitan areas.

In response to this health service delivery challenge, we developed the VIrtual Physiotherapist-led Evaluation of low back pain Referrals to spine surgeons (VIPER) model of care to help clinics identify new referrals who can be managed sooner by a physiotherapist and reduce wait times for those needing surgical review. Our intention is to conduct the first large-scale, multi-site randomised controlled trial of a model of care like VIPER in Australia and globally. However, prior to evaluating VIPER in a trial, we wanted to ensure VIPER was relevant and acceptable to end-users and scalable to improve Australian health services. As such we are co-designing VIPER with consumers, stakeholders (e.g., department heads, clinicians) and our partners from NSW health districts and professional and government organisations who were involved in developing our preliminary model of care.

The co-design of VIPER will be conducted across two phases: 1) qualitative interviews to explore stakeholder perspectives on VIPER (study reported here); and 2) co-design workshops to make sense of the information collected during the interviews in Phase 1, identify key insights and opportunities to VIPER, and generate creative solutions to refine the model ready for evaluation in a trial.^14^ The aim of this study (Phase 1) was to qualitatively explore patients, clinicians and other key stakeholders perceptions of our preliminary VIPER model of care.

## METHODS

### Study design and Ethics

This is an exploratory qualitative descriptive design study, involving in-depth semi-structured interviews exploring stakeholder perceptions about the preliminary VIPER model of care (Appendix 1). A qualitative descriptive design was selected for its utility in producing results aiming to present a straightforward, low inference account of key stakeholders’ views that stays close to the data and is written up so that the results can be of applied use. The COREQ checklist was used to ensure comprehensive reporting of this qualitative study.^15^ This project was approved by the University of Sydney Human Research Ethics Committee (HREC), project identifier 2024/HE001736.

### Participants

This study included adults living in Australia with a history of LBP (including those with lived experience of being referred to spine surgery clinics), AHPRA-registered clinicians who managed ≥5 people with LBP in the past year, and other key stakeholders working in government or healthcare organisations involved in LBP care. Participants were excluded if they did not live in Australia or did not speak English. Participants were sampled purposively for maximum variation in demographics, symptom presentation (patient-participants) and professional background and experience (clinician- and stakeholder-participants).

### Recruitment and data collection

Participants were recruited via social media advertisements, invitation emails to clinicians and key stakeholders within the authors’ professional networks, consumer organisations, and passive snowball sampling. Interested individuals first reviewed the participant information statement and provided informed consent before completing an online pre-interview questionnaire. For patients-participants, the pre-interview questionnaire captured demographics and LBP characteristics. For clinician-participants, it captured demographics, professional background, clinical experience, and experience managing LBP. For key stakeholder-participants, it captured demographics and professional background. Appendix 2 summarises these questionnaires.

### Semi-structured interviews

Purposive sampling was used to promote sample diversity based on pre-interview questionnaire responses. One-on-one semi-structured interviews were conducted via Zoom by three researchers (SM, JM and EW). SM is a physiotherapist who works in health policy and has extensive experience conducting qualitative interviews. JM and EW were medical students completing a capstone research project who began conducting interviews after receiving training from SM. The interview lasted between 30 to 60 minutes in duration. Interview guides, tailored to each stakeholder group, ensured consistency in the interview approach while allowing flexibility to explore emerging topics. Interview guides were mapped to the theoretical framework of acceptability of health interventions^16^ and the Consolidated Framework for Implementation Research (CFIR)^17^. Appendix 3 summarises these interview guides. Interviewers also presented participants with the schematic for the proposed VIPER model of care to help participants make sense of the questions that were asked (Appendix 1). Interview questions covered broad topics of experiences with LBP, experiences with the referral or referring to spine surgery clinics, and perceptions and acceptability of VIPER. Interviews were conducted until data saturation was achieved (i.e., no new perspectives or themes were identified after three consecutive interviews within each participant group). All audio interview recordings were uploaded onto OtterAI, an AI-powered transcription and note-taking tool that automatically converts spoken words into written text, to generate transcripts.

### Data analysis

Data analysis was conducted by two researchers (JM and EW) independently reviewing the transcripts (supervised by experience qualitative researchers SM and AH) and implementing an inductive descriptive analysis using NVivo. This involved open coding to generate codes, and then refining the codes into sub-themes and themes.^18^ JM and EW performed the analysis independently, then met together to harmonise their independently generated codes, sub-themes and themes. To enhance rigour and minimise personal bias, several members of the authorship team (SM, AH, TC, AG, JZ) also reviewed the coding of sub-themes themes and refined them following discussion with JM, EW and the broader authorship team.^19^

## RESULTS

### Participant characteristics

Recruitment occurred from May 2025 to October 2025. A total of 161 people completed the pre-interview questionnaires (61 people with LBP, 86 clinicians, and 14 key stakeholders). From these, we purposively sampled 39 adults to participate in interviews. The study sample included people with LBP (n=6), clinicians (n=26) including 16 physiotherapists, 5 spine surgeons, 2 pain specialist, 2 rheumatologist, and 1 registered nurse, and other key stakeholders (n=7). See Tables 1, 2, and 3 for participant characteristics, respectively. Recruitment of patient-participants was low relative to the other participant groups for several reasons. First, clinicians and key-stakeholders were interviewed first which meant data saturation was reached quicker when interviewing patient-participants. The preliminary VIPER model of care was developed with extensive consumer consultation, reflected by several consumers with lived experience of LBP and being referred to spine surgery clinics on our authorship team (ERG, EM, KT and DF).

**Table 1.** Characteristics of patient participants in the study.

| **Variables** | **Patient participants (n=6)** |
| --- | --- |
| Women, *n (%)* | 4 (66.6) |
| Age, years, *mean (range)* | 51.8 (30 to 80) |
| Education, *n (%)* |  |
| *Advanced diploma or diploma* | 1 (16.7) |
| *Bachelor degree or higher* | 5 (83.3) |
| Household income bracket, \$/year, *n (%)* | |
| *\$1 - \$41,599* | 1 (16.7) |
| *\$41,600 - \$83,199* | 4 (66.6) |
| *\$83,200 - \$156,999* | 0 (0.0) |
| *\$156,000 - \$311,999* | 1 (16.7) |
| Private healthcare insurance, *n (%)* |  |
| *Yes* | 4 (66.6) |
| *No* | 2 (33.4) |
| Healthcare LBP intervention, *n (%)* |  |
| *Seen surgeon* | 5 (83.3) |
| *Had surgery* | 1 (16.7) |

**Table 2.**
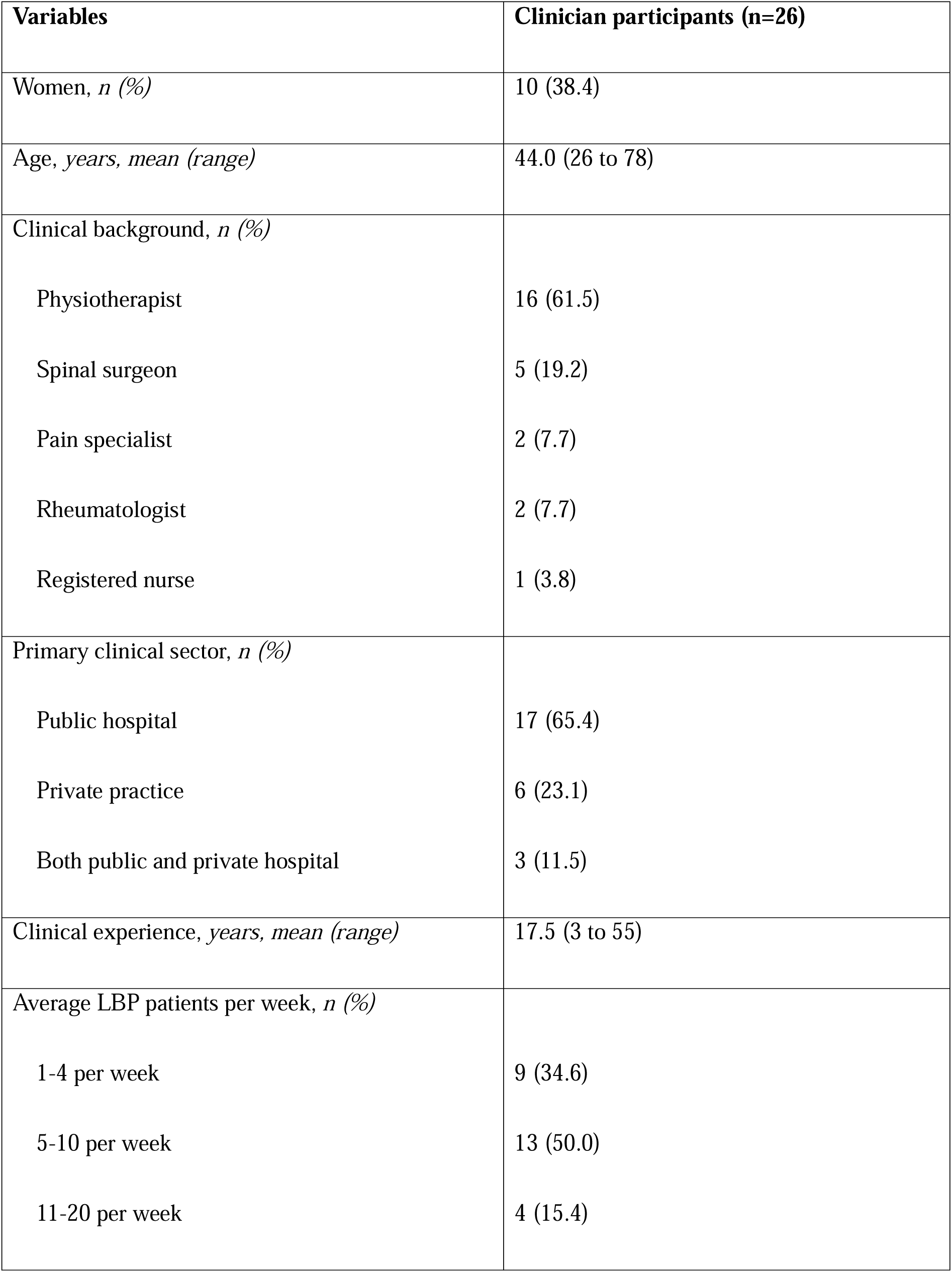
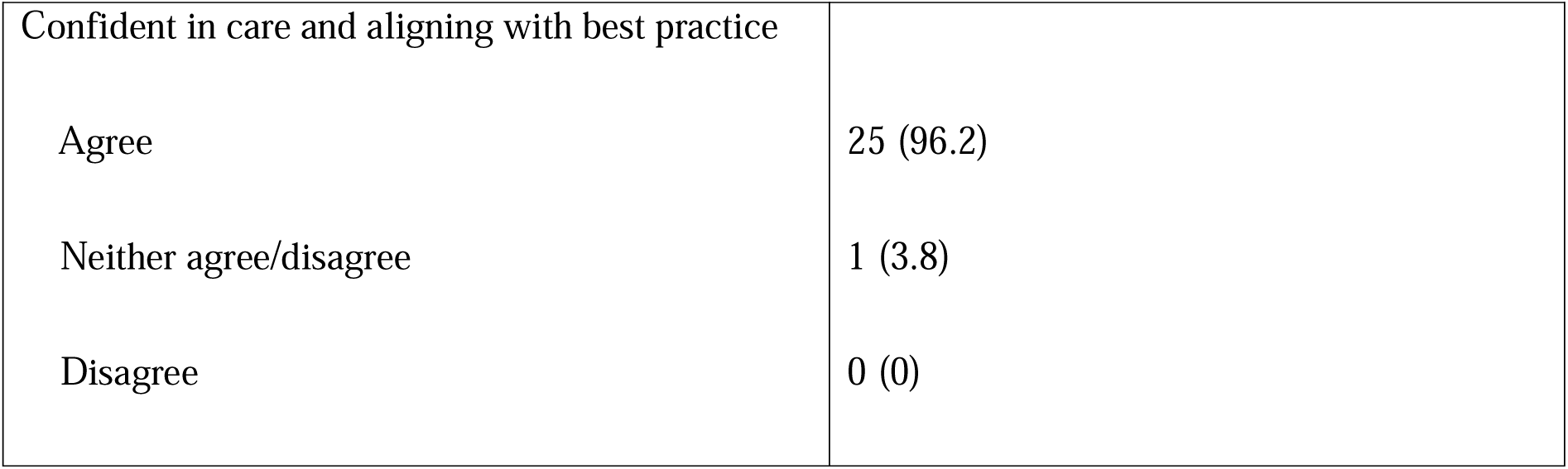
Characteristics of clinician participant.

| **Variables** | **Clinician participants (n=26)** |
| --- | --- |
| Women, *n (%)* | 10 (38.4) |
| Age, years, *mean (range)* | 44.0 (26 to 78) |
| Clinical background, *n (%)* |  |
| Physiotherapist | 16 (61.5) |
| Spinal surgeon | 5 (19.2) |
| Pain specialist | 2 (7.7) |
| Rheumatologist | 2 (7.7) |
| Registered nurse | 1 (3.8) |
| Primary clinical sector, *n (%)* |  |
| Public hospital | 17 (65.4) |
| Private practice | 6 (23.1) |
| Both public and private hospital | 3 (11.5) |
| Clinical experience, years, *mean (range)* | 17.5 (3 to 55) |
| Average LBP patients per week, *n (%)* |  |
| 1-4 per week | 9 (34.6) |
| 5-10 per week | 13 (50.0) |
| 11-20 per week | 4 (15.4) |
| Confident in care and aligning with best practice |  |
| Agree | 25 (96.2) |
| Neither agree/disagree | 1 (3.8) |
| Disagree | 0 (0) |

**Table 3.** Characteristics of other key stakeholder participants.

| **Variables** | **Other key stakeholder participants (n=7)** |
| --- | --- |
| Women, *n (%)* | 3 (42.8) |
| Age, years, *mean (range)* | 44.1 (33 to 53) |
| Stakeholder type, *n (%)* |  |
| Healthcare governmental organisation | 7 (100) |
| Position, *n (%)* |  |
| Director | 1 (14.3) |
| Head of department | 1 (14.3) |
| Deputy head | 2 (28.6) |
| Manager | 2 (28.6) |
| Team leader | 1 (14.3) |

### Semi-structured interviews

Four main themes were identified, each comprising 3 to 4 subthemes. Collectively, these findings offer insight into stakeholders’ perceptions and highlight barriers and enablers to implementing VIPER for adults with LBP referred to public hospital spine surgery clinics.

### *Theme 1*. Gaps in current care pathways and implementation considerations

Participants consistently described gaps in current LBP care pathways that compromise timely, effective management, and highlighted key considerations for service design and implementation. Illustrative quotes for subthemes 1.1 - 1.4 are presented in Table 4.

**Table 4:**
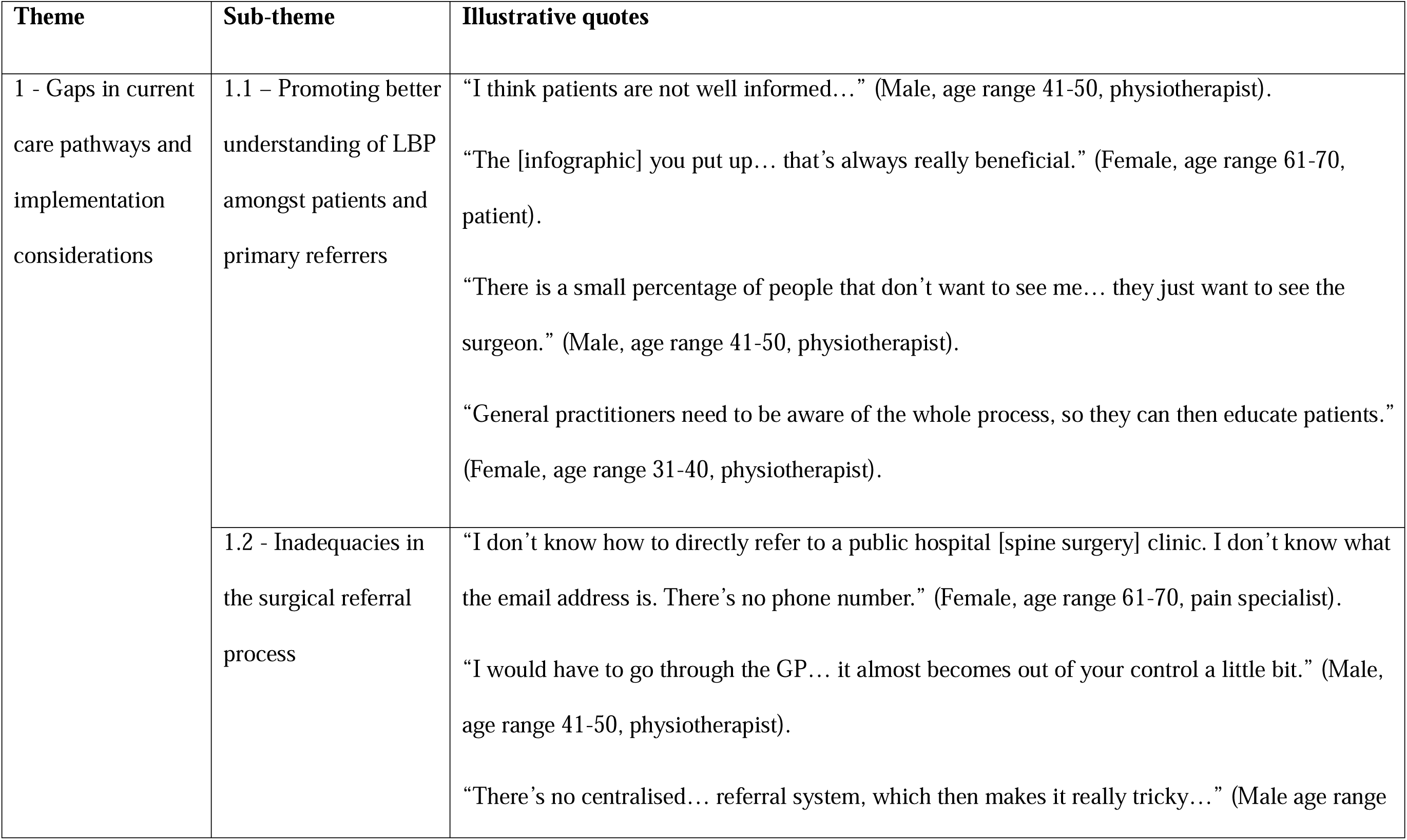
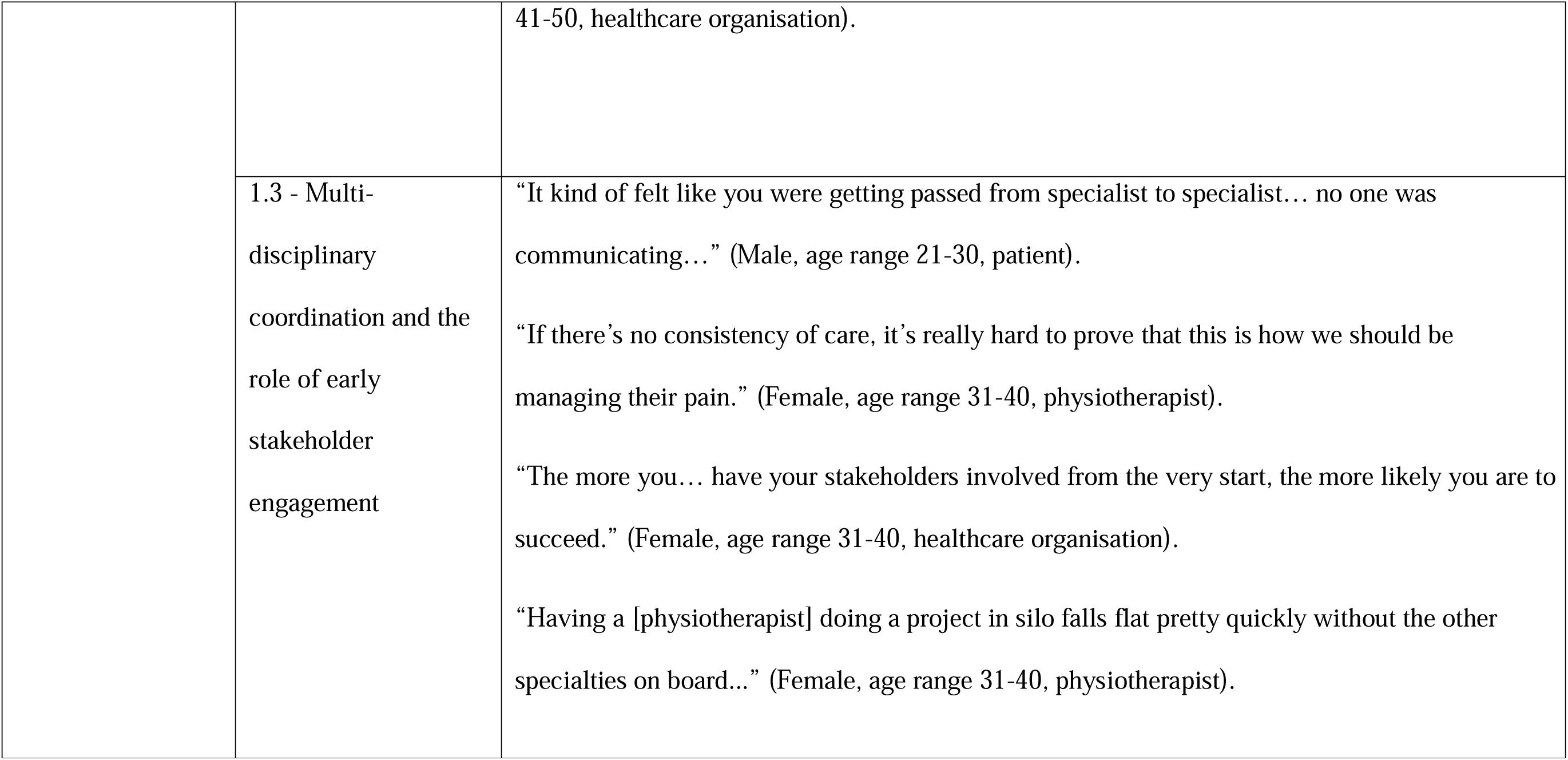
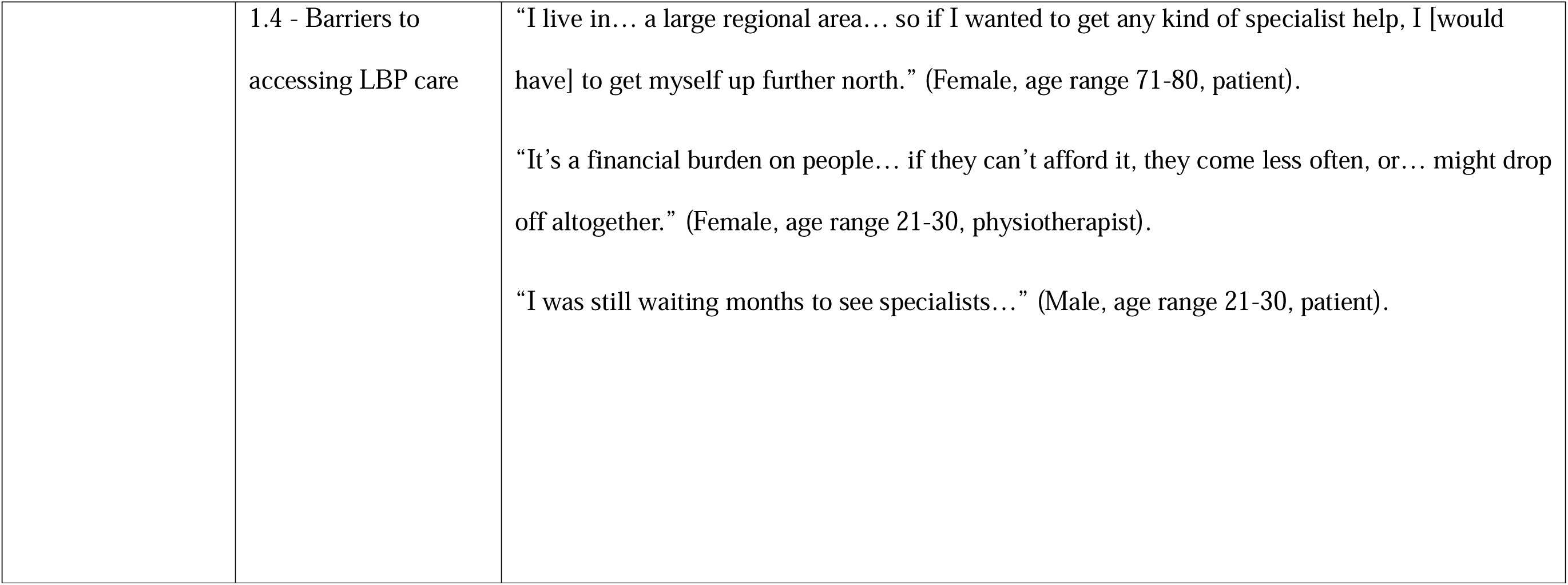
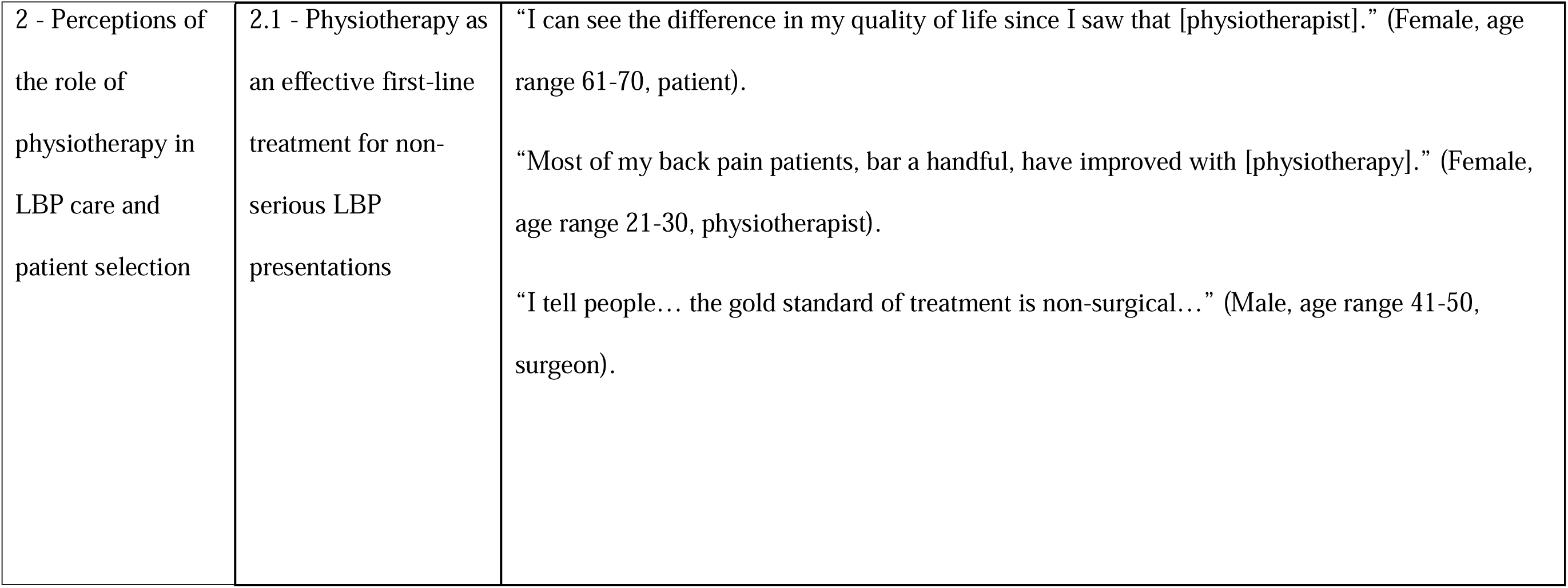
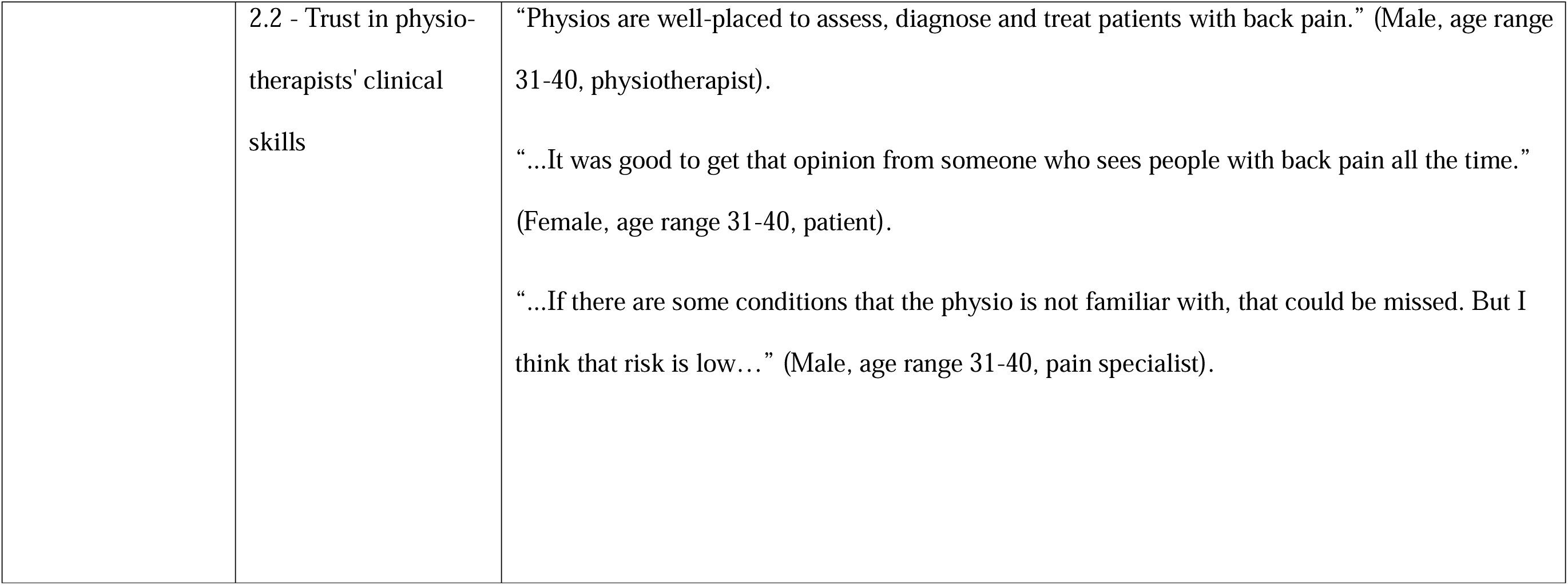
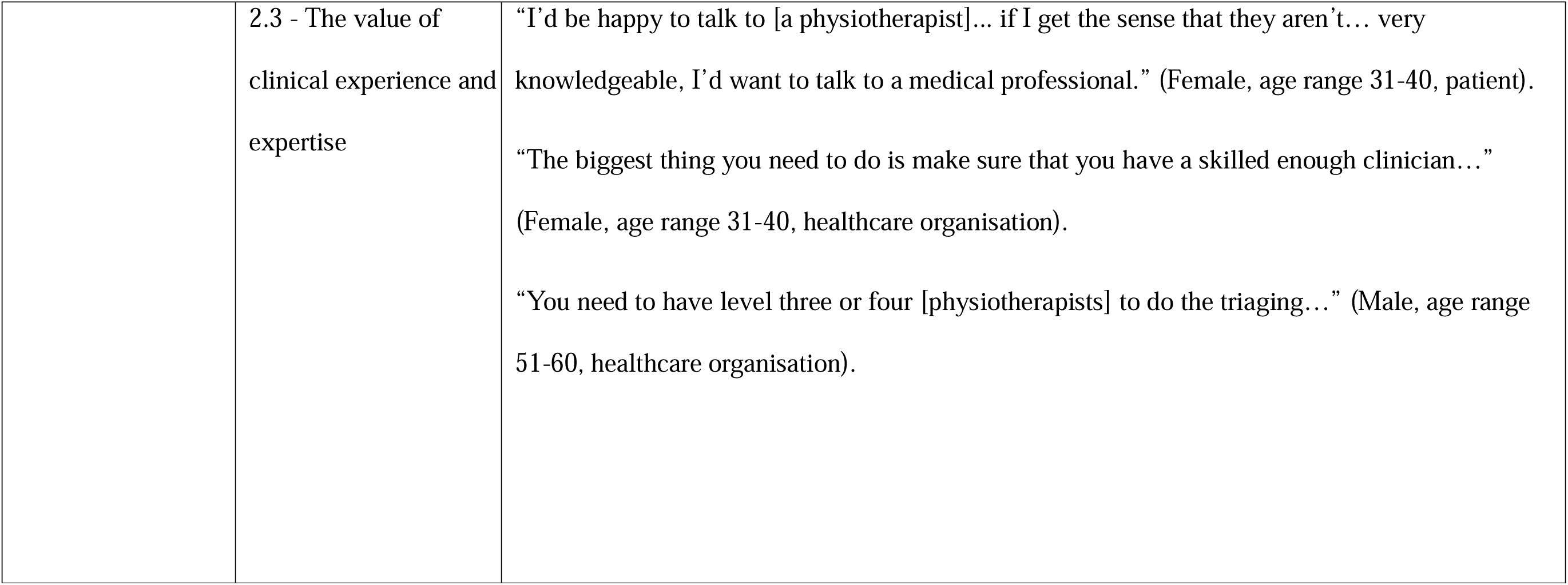
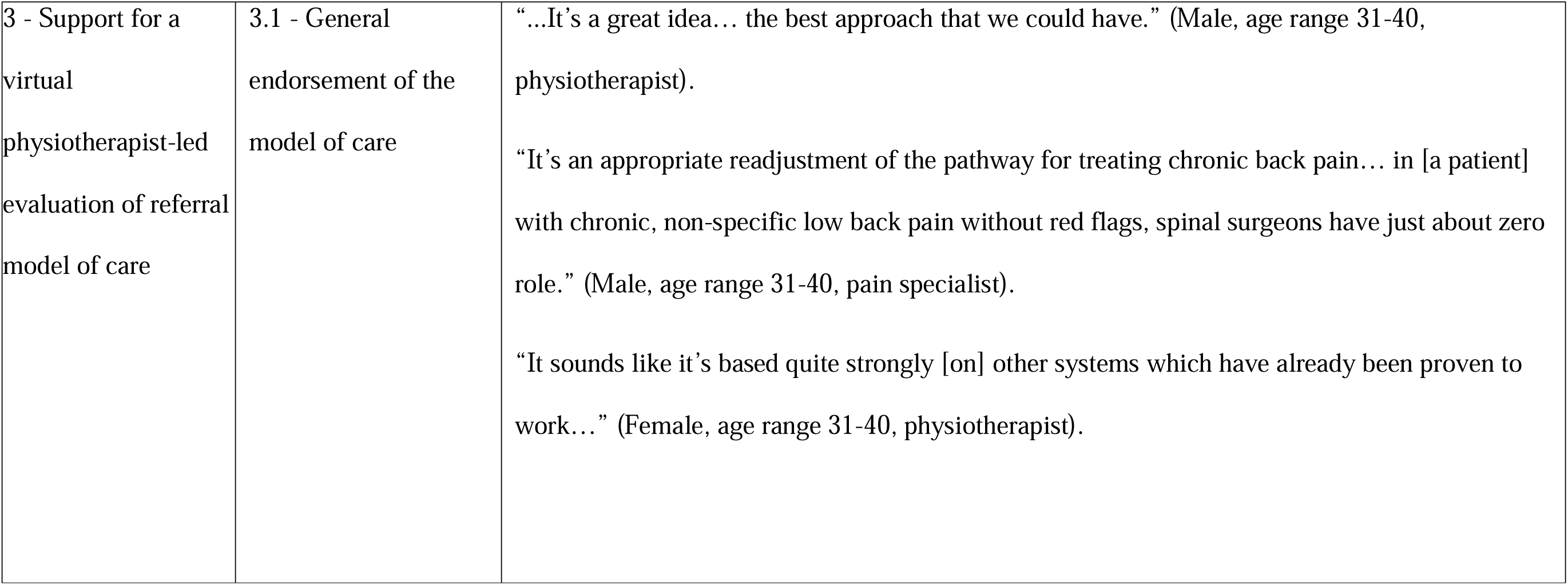
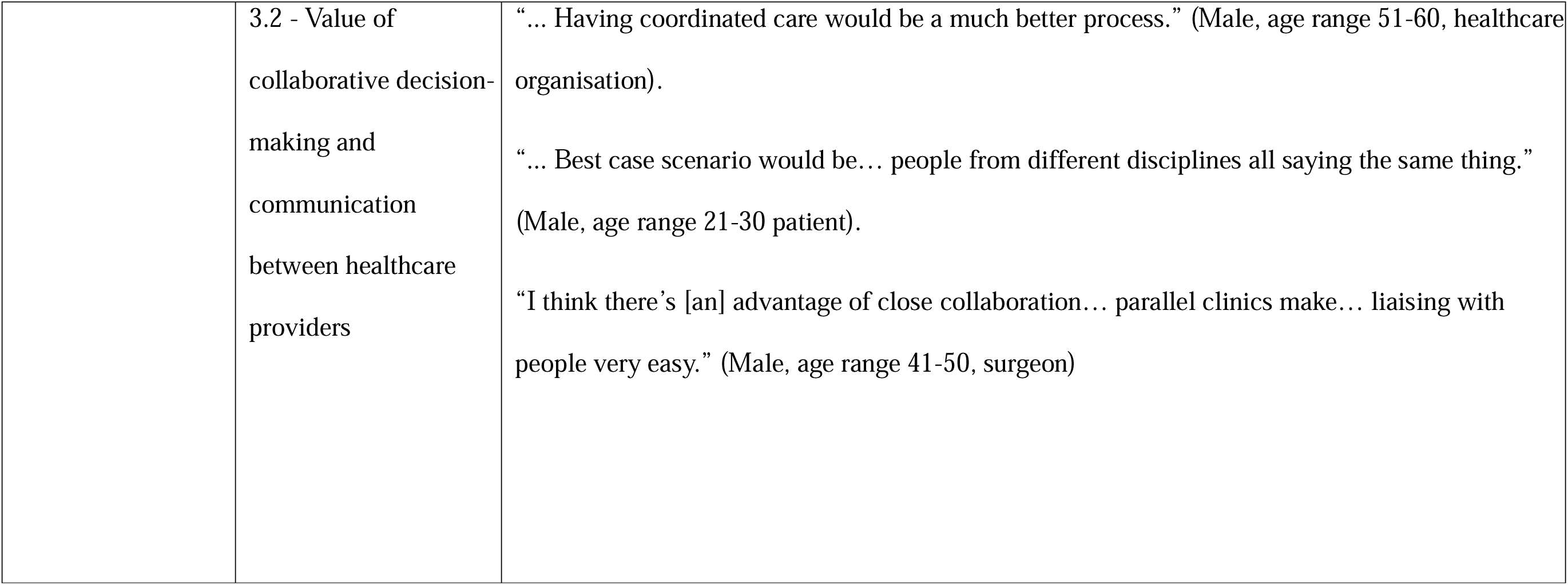
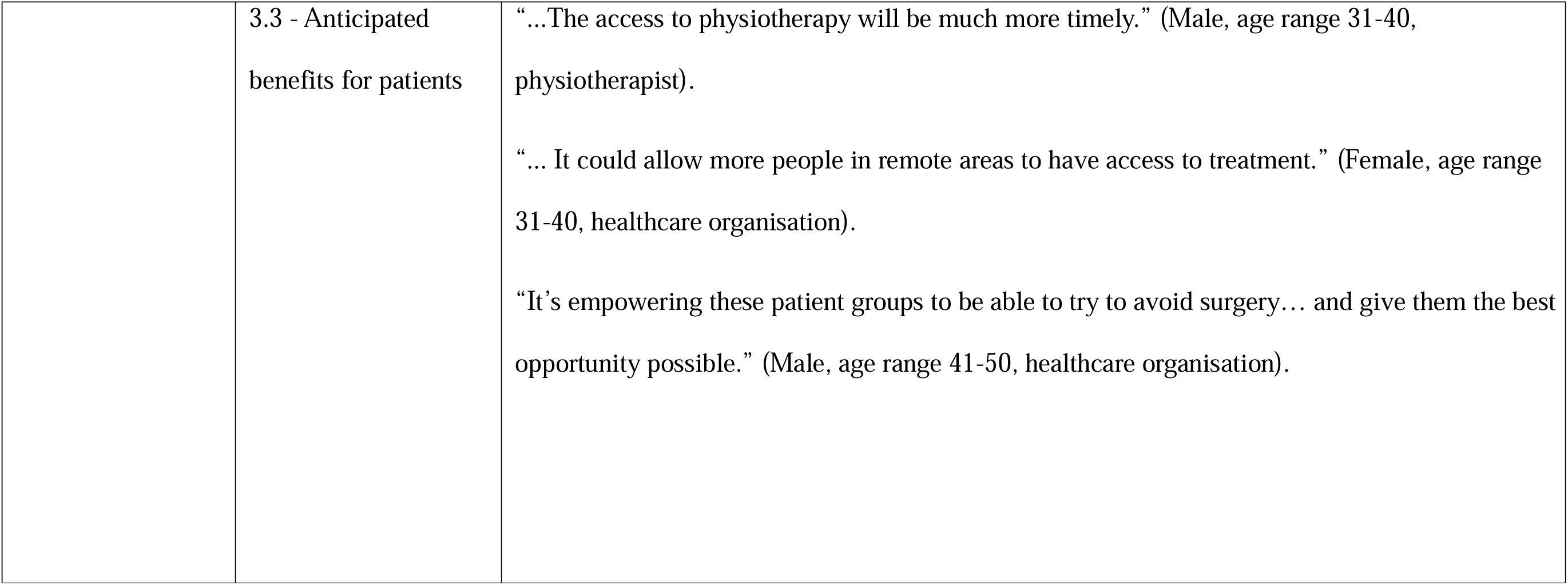
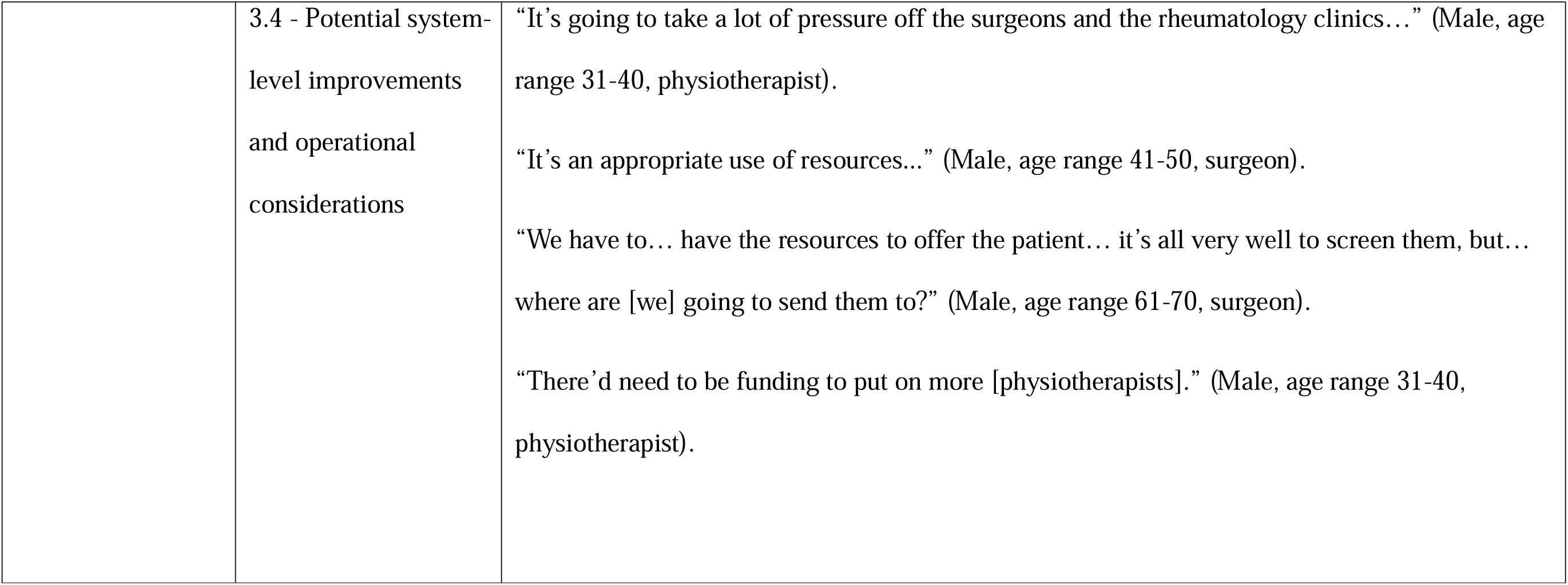
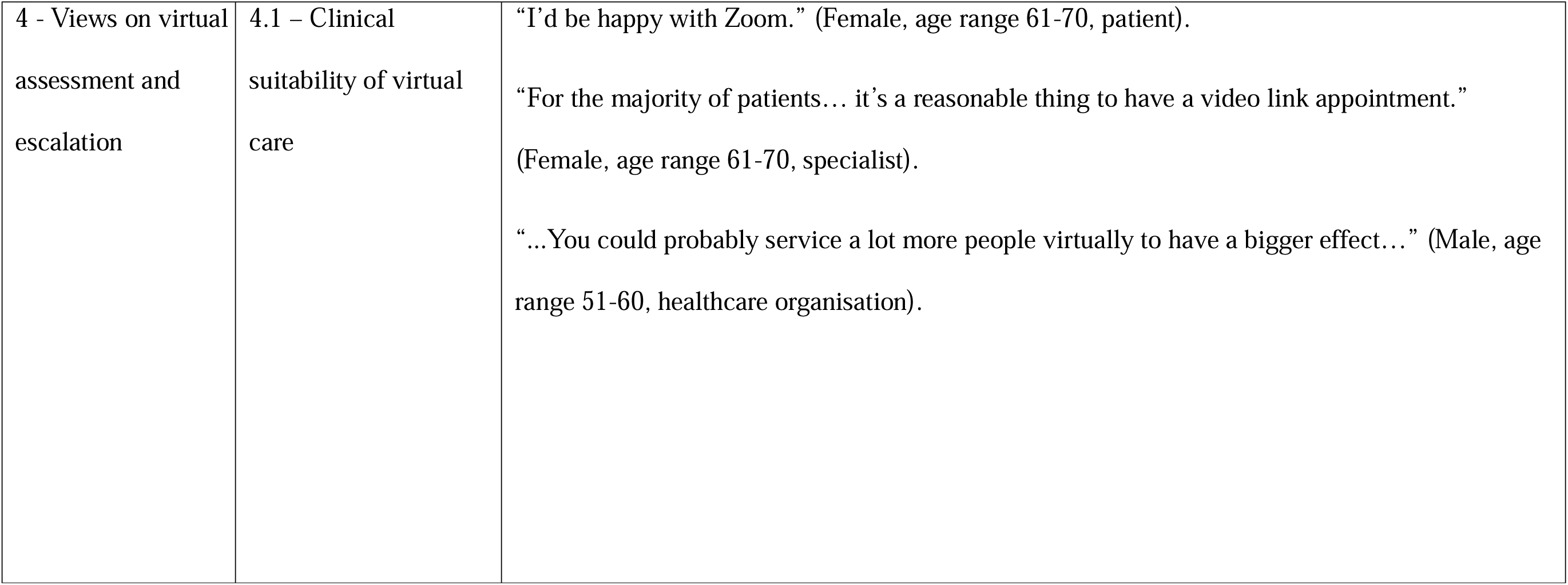
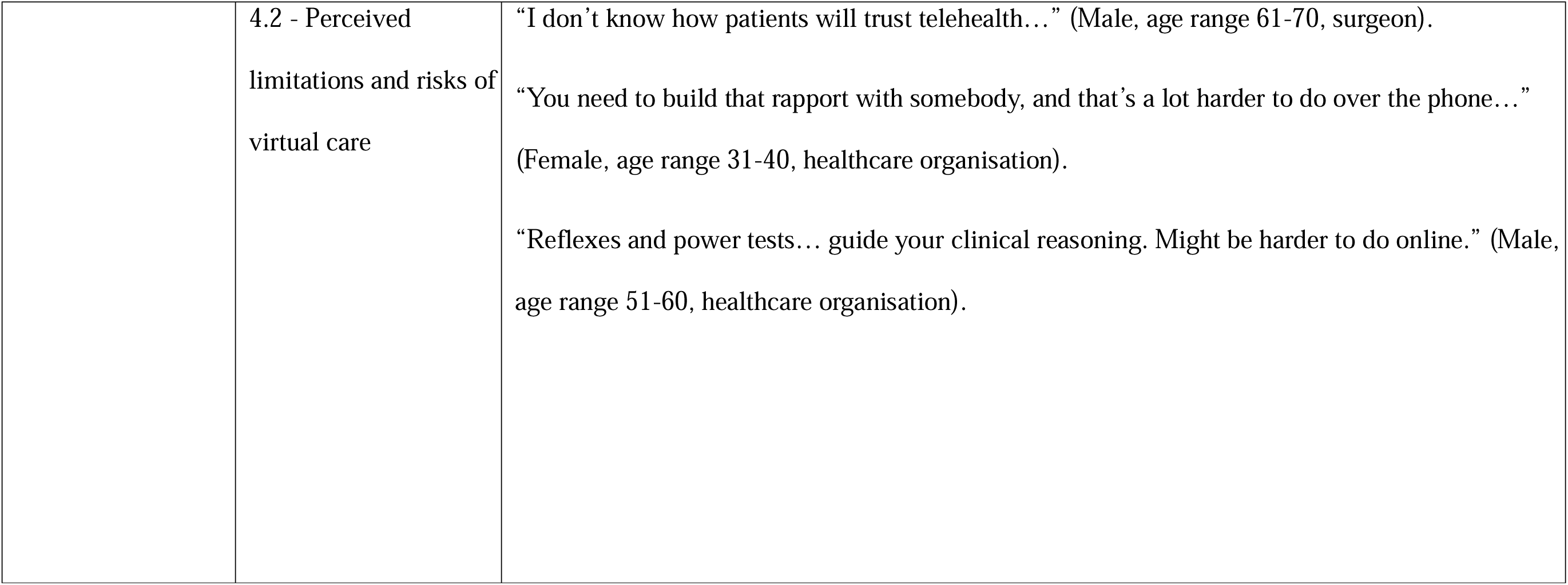
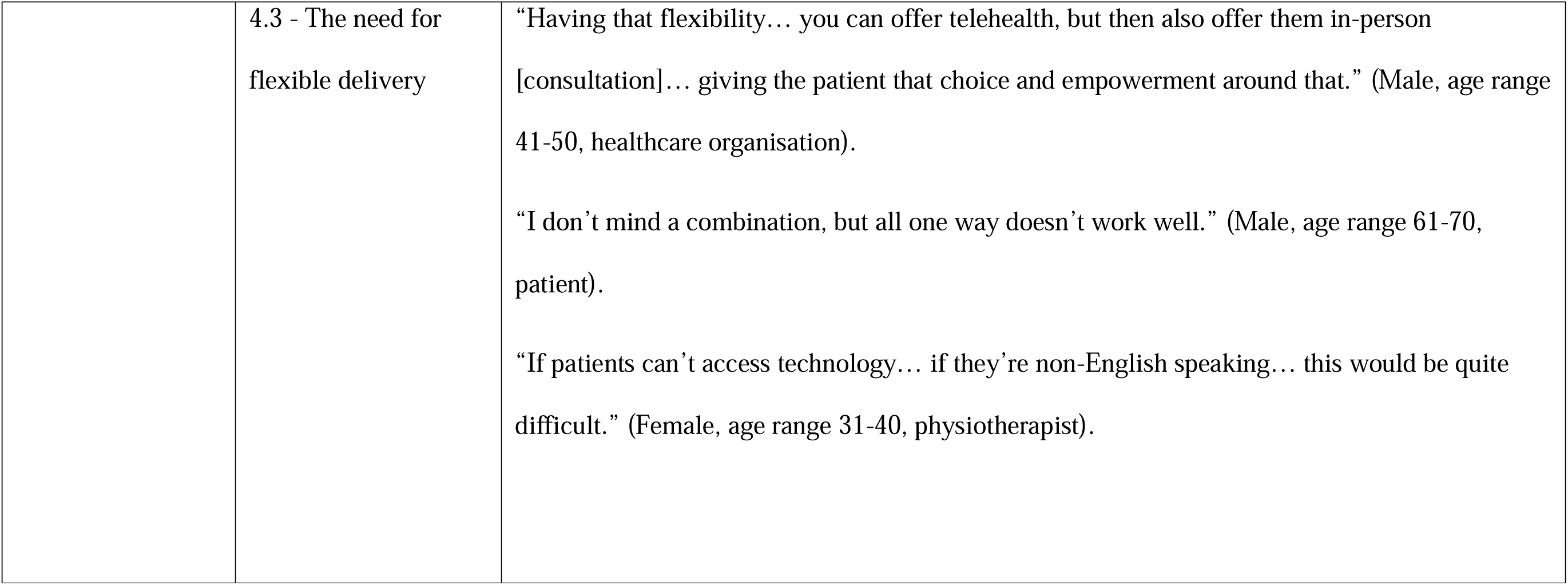
Summary of themes, subthemes and illustrative quotes from stakeholder interviews.

#### Subtheme 1.1: Promoting better understanding of LBP amongst patients and primary referrers

All stakeholder groups identified education as being integral to effective LBP management. Clinicians flagged limited patient understanding of LBP, which leads to fear-avoidance behaviours (e.g. kinesiophobia) and unrealistic expectations (e.g. viewing surgery as a definitive solution for LBP). Patients echoed these concerns, reporting insufficient information and noting that earlier, comprehensive education could have improved both physical and psychological outcomes.

> “I think patients are not well informed…” (Male, age range 41-50, physiotherapist)

Participants stressed the significance of educating patients on VIPER’s purpose, scope and benefits, for example, through infographics or short videos. Setting expectations early (i.e. clarifying that patients would initially see a physiotherapist) was deemed important to avoid confusion or discontent. Improving awareness about VIPER among general practitioners (GPs) was also considered critical to support integration into existing workflows and referral pathways.

#### Subtheme 1.2: Inadequacies in the surgical referral process

Patients described the surgical referral process as slow, confusing and anxiety-inducing. Clinicians criticised it as inconsistent, lacking transparency, and difficult to navigate. Some physiotherapists expressed frustration that, despite their frontline role in LBP care, referrals must be made by GPs. These inadequacies were perceived to delay treatment. Several participants advocated for a centralised, standardised referral system to improve access and consistency.

> “There’s no centralised… referral system, which then makes it really tricky…” (Male, age range 41-50, healthcare organisation)

#### Subtheme 1.3: Multidisciplinary coordination and the role of early stakeholder engagement

Poor communication between physiotherapists, GPs, and surgeons was seen as a barrier to coordinated care. Inconsistent messaging was believed to confuse patients, foster misconceptions, and complicate management. Some patients noted that conflicting advice undermined their trust. Collaboration and shared decision-making were seen as essential for promoting continuity and confidence in care.

> “It kind of felt like you were getting passed from specialist to specialist… no one was communicating…” (Male, age range 21-30, patient)

To support coordinated service delivery, participants highlighted the need to engage key stakeholders (e.g. GPs, surgeons, and executives) early in the design and implementation process. This was viewed as critical for achieving buy-in, as some felt that bottom-up approaches might be insufficient without strong support from medical professionals, whose advocacy was considered key to enhancing the VIPER model’s credibility and uptake. However, gaining surgeon support was acknowledged as a potential challenge, particularly if the model of care is perceived to reduce surgical caseloads.

#### Subtheme 1.4: Barriers to accessing LBP care

Access barriers were frequently reported. Challenges were noted for rural or remote residents, where local services are often limited and travel and accommodation costs are high. Affordability and long wait times were identified as key barriers, with many unable to afford private healthcare, subsequently facing long waits in the public system. Transport and mobility issues, particularly among those with chronic pain or disability, were thought to further limit access.

> “I live in… a large regional area… so if I want to get any kind of specialist help, I [need] to get myself further north.” (Female, age range 71-80, patient)

Collectively, the findings from Theme 1 reveal significant gaps in current LBP care pathways, underscoring the need for improved education, communication, care coordination and equitable care access.

### Theme 2: Perceptions of the role of physiotherapy in LBP care and patient selection

Physiotherapy was endorsed as a central component of LBP care, particularly in the management of non-specific LBP and stable and mild radiculopathy, reinforcing physiotherapists’ role in the triage and management of most patients with LBP. Illustrative quotes for subthemes 2.1 - 2.3 are presented in Table 4.

#### Subtheme 2.1: Physiotherapy as an effective first-line treatment for non-serious LBP presentations

Patients consistently reported positive experiences with physiotherapy, noting improvements in physical function and quality of life. Across all stakeholder groups, physiotherapy interventions were viewed as appropriate and effective for LBP, especially before surgical consideration and intervention.

> “Most of my back pain patients… have improved with [physiotherapy].” (Female, age range 21-30, physiotherapist)

#### Subtheme 2.2: Trust in physiotherapists’ clinical skills

All stakeholders expressed confidence in physiotherapists’ ability to triage and manage most LBP cases, perceiving them as suitable first-contact practitioners in surgical triage pathways. Some participants noted a small risk of diagnostic oversight, though this was not considered a major limitation.

> “…It was good to get that opinion from someone who sees people with back pain all the time.” (Female, age range 31-40, patient)

#### Subtheme 2.3: The value of clinical experience and expertise

Support for VIPER was contingent on the triaging physiotherapist having adequate clinical experience; the safety and success of VIPER was perceived to depend heavily on this. Definitions of “adequate” varied, though some suggested the role be filled by Level 4 Physiotherapists (NSW Health Staff Award Rate) or those with advanced training in the management of LBP. Other desirable attributes included experience managing chronic pain, strong ability to identify surgical indications, and confidence using virtual care platforms.

> “The biggest thing you need to do is make sure that you have a skilled enough clinician…” (Female, age range 31-40, healthcare organisation)

The findings of Theme 2 highlight a strong belief that physiotherapists play a key role in LBP care and emphasise the importance of clinical expertise in advanced practice roles.

### Theme 3: Support for a virtual physiotherapist-led evaluation of referral model of care

Participants broadly supported the VIPER model of care, recognising its potential to address inefficiencies in current care pathways. Illustrative quotes for subthemes 3.1 - 3.4 are presented in Table 4.

#### Subtheme 3.1: General endorsement of the model of care

All participants supported VIPER, viewing it as a timely response to rising demand and system inefficiencies such as long wait times for surgical review. It was perceived to offer more streamlined, proactive care and reduce the need to see a spine surgeon in many cases. Anecdotal reports of similar services, operating successfully in other parts of the country, reinforced confidence in the model of care.

> “…It’s a great idea… the best approach that we could have.” (Male, age range 31-40, physiotherapist)

#### Subtheme 3.2: Value of collaborative decision-making and communication between healthcare providers

Close collaboration between physiotherapists and surgeons was considered a strength of the VIPER model of care. Clinicians believed it would enhance safety, and patients noted that having multiple providers aligned in care decisions would increase their satisfaction. However, some suggested that, over time, strong trust between the clinicians could eliminate the need to discuss every case, allowing physiotherapists to practice with greater independence. Co-location or parallel clinics were proposed to support prompt communication and escalation of care where needed.

> “I think there’s [an] advantage of close collaboration… parallel clinics make… liaising with people very easy.” (Male, age range 41-50, surgeon)

#### Subtheme 3.3: Anticipated benefits for patients

Earlier access to physiotherapy (and other non-surgical care options) was identified as the main benefit of this model of care for patients. Patient-participants felt that timely physiotherapy intervention could improve outcomes and potentially reduce the need for surgery.

> “… The access to physiotherapy will be much more timely.” (Male, age range 31-40, physiotherapist)

#### Subtheme 3.4: Potential system-level improvements and operational considerations

The VIPER model of care was seen as a strategy to improve system efficiency by reducing wait times, alleviating pressure on spine surgery clinics, and promoting efficient resource use, with potential cost savings from reduced imaging, consultations and surgeries. However, clinician- and key stakeholder-participants highlighted challenges related to funding, workforce shortages and limited resources. Without adequate staffing and support, these participants noted that physiotherapy clinics risk being overwhelmed, potentially compromising quality of care. Careful planning was encouraged to ensure sustainable, high-quality service delivery.

> “We have to… have the resources to offer the patient… It’s all very well to screen them, but… where are [we] going to send them to?” (Male, age range 61-70, surgeon)

This quote directly supports the need to fund a physiotherapist to intercept referrals in spine surgery clinics, rather than the physiotherapists operating in their capacity as an outpatient physiotherapist; a setting which also has issues with long wait times.

The findings of Theme 3 highlight strong stakeholder support, suggesting that VIPER is acceptable and aligns with system-level needs, provided it is underpinned by appropriate resourcing.

### Theme 4: Views on virtual assessment and escalation

Participants acknowledged the benefits of virtual assessment and management but emphasised the importance of flexible delivery to accommodate patients’ preferences or clinical complexity.

Illustrative quotes for subthemes 4.1 - 4.3 are presented in Table 4.

#### Subtheme 4.1: Clinical suitability of virtual care

Virtual care (e.g., telehealth, videoconference appointment) was generally viewed as appropriate for triaging and managing most LBP presentations such as non-specific LBP and stable or mild radiculopathy, regardless of pain intensity. It was considered efficient for screening large volumes of referrals and providing timely care, especially for people in rural, remote and regional areas. Most clinician-participants agreed that history-taking, education and exercise prescription are feasible to be delivered virtually. Virtual care was thought to be particularly beneficial for those with mobility limitations, chronic pain, or living in geographically isolated areas.

> “For the majority of patients… It’s a reasonable thing to have a video link appointment.” (Female, age range 61-70, pain specialist)

Clinicians and stakeholders viewed virtual assessment as feasible but emphasised the need for escalation to an in-person assessment or surgeon review if written referrals missed important surgical indications (e.g., severe or progressive radiculopathy which would need to be assessed in-person) or symptoms were worsening.

#### Subtheme 4.2: Perceived limitations and risks of virtual care

While generally supportive of virtual care, participants across all groups acknowledged its limitations. Many cited difficulties in building trust and rapport virtually, and the reduced capacity for physical assessment (e.g. neurological exams or subtle clinical signs), raising some concerns about diagnostic accuracy. As a result, most participants expressed a preference for in-person care but acknowledged having the option of virtual assessment and/or management would benefit many patients.

> “Reflexes and power tests… guide your clinical reasoning. Might be harder to do online.” (Male, age range 51-60, healthcare organisation)

#### Subtheme 4.3: The need for flexible delivery

A flexible delivery approach (offering patients the choice of virtual or in-person care) was frequently suggested. Participants highlighted that exclusive reliance on virtual care may disadvantage certain populations, such as those without internet access or non-English speakers. Clinicians noted that some patients, such as those with complex conditions, may require in-person evaluation.

> “I don’t mind a combination, but all one way doesn’t work well.” (Male, age range 61-70, patient)

These findings highlight that while virtual care can enhance access and efficiency, flexible delivery is needed to ensure care remains equitable and clinically appropriate.

## DISCUSSION

This qualitative study identified diverse but overall supportive perceptions of VIPER from people with LBP, clinicians and key stakeholders. All participants broadly supported VIPER as a timely, pragmatic response to system constraints, including long wait times, inconsistent referral pathways, and suboptimal use of specialist resources. Four themes emerged: 1) current gaps in LBP care pathways and implementation considerations (covering need for appropriate patient and clinician education, referral inefficiencies, coordination challenges, and access barriers); 2) perceptions of the role of physiotherapy in LBP care and patient selection for VIPER; 3) support for VIPER as a means to improve patient outcomes and health system efficiency; and 4) views on virtual assessment and escalation, recognising the value of hybrid models and its limitations. These themes will be discussed in subsequent co-design workshops (Phase 2 of the VIPER co-design process) to identify key insights and opportunities to VIPER and generate creative solutions to refine the model ready for evaluation in a large, multisite randomised controlled trial.^14^

Consistent with previous research, participants identified significant gaps in current LBP care pathways that compromise timely, effective management. Inadequate patient education was frequently noted, contributing to fear-avoidance behaviours, which are known to worsen outcomes.^20^ Clinical guidelines for the management of LBP^21^ including from NICE^22^ emphasise education and reassurance as foundational to LBP management, with evidence showing that even simple educational interventions can reduce fear-avoidance beliefs and support recovery.^23^ ^24^ As such, embedding structured education within the VIPER model of care will be critical to optimise outcomes and satisfaction. The existing surgical referral process in many spine surgery clinics was described as slow, confusing, and inconsistent, echoing calls to reconfigure, centralise or standardise referral systems to improve access and equity.^25–27^ Poor communication between physiotherapists, GPs and surgeons fractures coordination of care^28^ and undermines patients’ trust,^29^ highlighting the importance of early, ongoing engagement with relevant stakeholders to not only strengthen care^30^ but also foster buy-in. The reports of access barriers reflect known inequities in musculoskeletal care,^31–33^ particularly for rural or remote populations,^34^ ^35^ reinforcing the need for equity-focused models of care consistent with broader health equity principles.^36^

Physiotherapists were strongly endorsed as able to deliver effective first-line treatment for LBP and be effective triage clinicians, aligning with evidence that non-surgical care for LBP offers comparable outcomes with fewer adverse events to more invasive interventions.^2^ ^37^ ^38^ Patient and surgeon confidence in physiotherapists’ diagnostic ability supports their role as triage clinicians, consistent with evidence that physiotherapists can accurately diagnose and manage most LBP cases.^39^ Sufficient clinical expertise was viewed as essential for maintaining quality and safety; this is supported by research indicating that physiotherapists’ diagnostic accuracy can match that of orthopaedic surgeons.^40^ Still, where physiotherapists lead triage services, credentialing, clear governance and a well-defined scope of practice may be important to sustain care quality and interprofessional confidence. Evaluations of similar physiotherapist-led services demonstrate high patient and referrer satisfaction,^12^ ^41–43^ and shorter wait times,^42^ ^44^ ^45^ without compromising clinical outcomes compared to surgeon-led care.^12^ ^40^ However, without adequate resourcing for follow-up conservative care, triage alone risks creating bottlenecks in care pathways,^42^ ^46^ highlighting the need for coordinated planning to ensure sustainability and quality.^47^ For example, funding a physiotherapist to work in spine surgery clinics to perform both the assessment and management of patients with LBP was raised as vital to the success of VIPER.

Virtual assessment was seen as appropriate and efficient for triaging and managing most LBP presentations (especially where history-taking, education and exercise prescription are central). This mirrors the findings of a study exploring telehealth in musculoskeletal care, in which it was perceived to be appropriate for less complex cases, and valuable for improving access for rural and remote patients.^48^ Participants highlighted limitations in physical examination, diagnostic certainty and rapport-building (issues also highlighted in broader research,^49^ ^50^ prompting suggestions of a hybrid VIPER model where participants could choose to receive their assessment and/or treatment virtually or in-person. Evidence regarding the comparability of virtual care to in-person care is still in its infancy for musculoskeletal conditions relative to the evidence-based of in-person care, though many studies have found positive outcomes for virtual physiotherapy,^51^ ^52^ with an umbrella review concluding that virtual care offers non-inferior outcomes for musculoskeletal care while enhancing care access.^53^ Virtual care is likely best-placed in a hybrid model, supporting patient choice and allowing in-person assessment when required.^54^

### Strengths and limitations

This study’s strengths include engagement of 39 participants across three stakeholder groups, offering diverse perspectives and insights specific to the Australian healthcare context, where evidence for such a virtual triage model remains scarce. The co-design component of this work enhances this study’s relevance, while iterative thematic analysis and independent coding strengthened analytical rigour. Limitations include potential selection bias, as stakeholders self-selected to participate and likely had stronger opinions on LBP care than the wider population. Over representation of physiotherapists (n=16) and underrepresentation of surgeons (n=5) may have influenced thematic emphasis towards certain viewpoints. However, this likely reflects the relative number of physiotherapists compared to spine surgeons in Australia. The relatively small numbers of patients (n=6) may be seen as a limitation. However, clinicians and key-stakeholders were interviewed first which meant data saturation was reached quicker when interviewing patient-participants. The preliminary VIPER model of care was also developed with extensive consumer consultation, reflected by several consumers with lived experience of LBP and being referred to spine surgery clinics on our authorship team (ERG, EM, KT and DF). This meant we needed to recruit less people with LBP to this study as their views were largely captured from our consumer advisory panel. It should be noted that, as the model of care has not yet been implemented, participants’ views in this study are largely hypothetical and may differ in practice.

### Implications for practice

Broad support from patients, clinicians and key stakeholders provides a solid foundation for further refining VIPER in co-design workshops, with the intention to evaluate a more refined model of care in a large, multi-site trial conducted across metropolitan, regional, rural and remote parts of Australia. Suggested elements to improve the success of VIPER include clear escalation from physiotherapist triage to surgical review, collaborative decision-making and communication between physiotherapists and spine surgeons, clear patient selection largely targeting non-serious LBP (e.g., non-specific LBP, mild and stable radiculopathy), ongoing stakeholder engagement, comprehensive patient and GP education, sufficient resourcing of follow-up conservative care, and a flexible hybrid delivery model accommodating patient preferences and clinical needs. These findings add to the evidence supporting integration of advanced physiotherapy roles and virtual care delivery for LBP, presenting a model that could be extended to other musculoskeletal conditions within the Australian healthcare system and beyond if later shown to be effective in a randomised controlled trial.

### Conclusion

Stakeholders viewed our preliminary VIPER model of care as feasible, acceptable and well-positioned to address current inefficiencies in LBP care. This study underscores the important role that physiotherapists can play in improving access to care for LBP, reducing spine surgery clinic wait times, and promoting conservative non-surgical management aligned with best practice guidelines. Integrating virtual healthcare delivery into VIPER offers a flexible, patient-centred approach with potential to enhance equity and system efficiency. These findings provide a strong foundation for further refining VIPER in co-design workshops, with the intention to evaluate a more refined model of care in a large, multi-site trial.

## Data Availability

All data produced in the present study are available upon reasonable request to the authors.

---

**Appendix 1:**
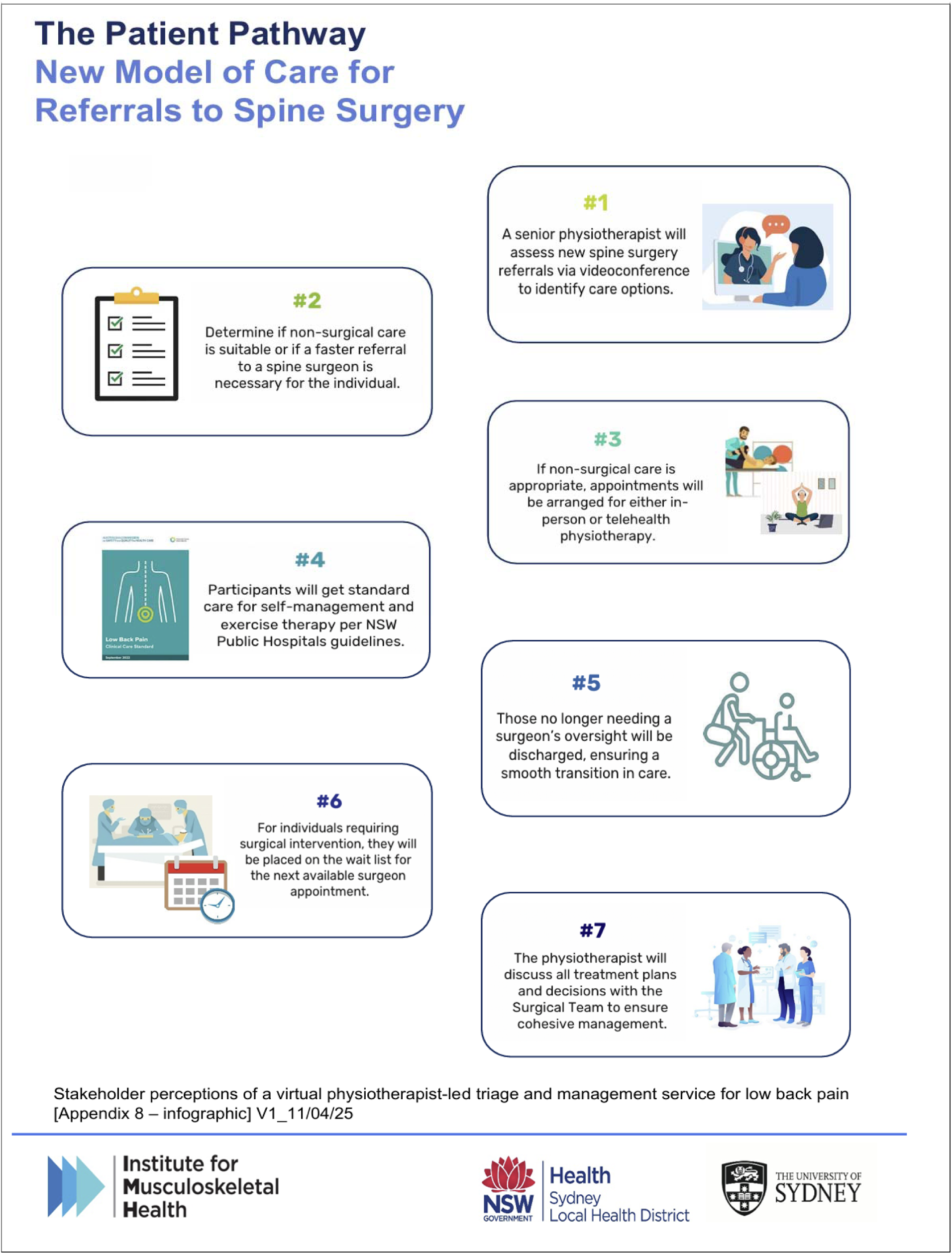
Schematic for the proposed model-of-care.

**Appendix 2:**
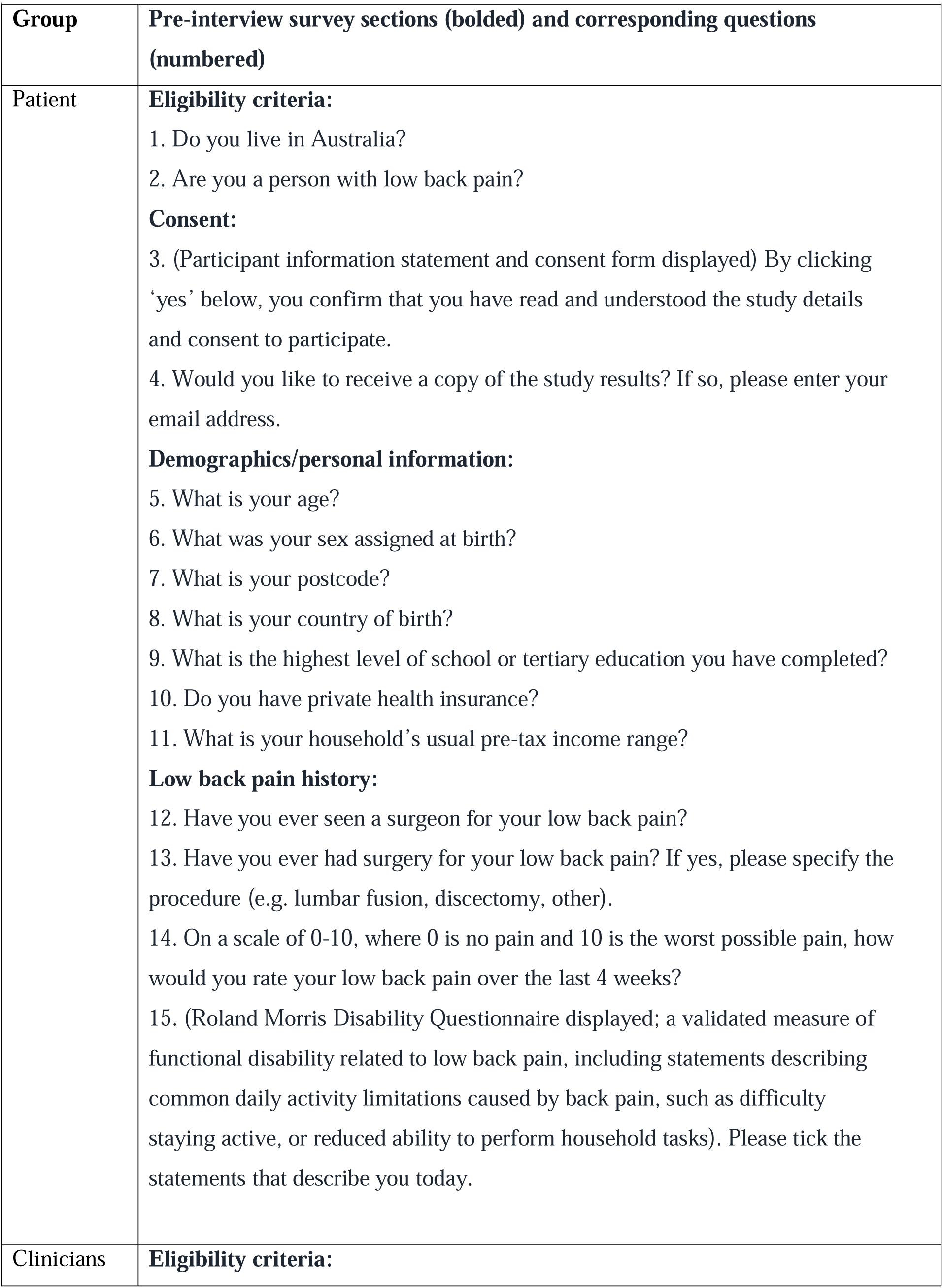
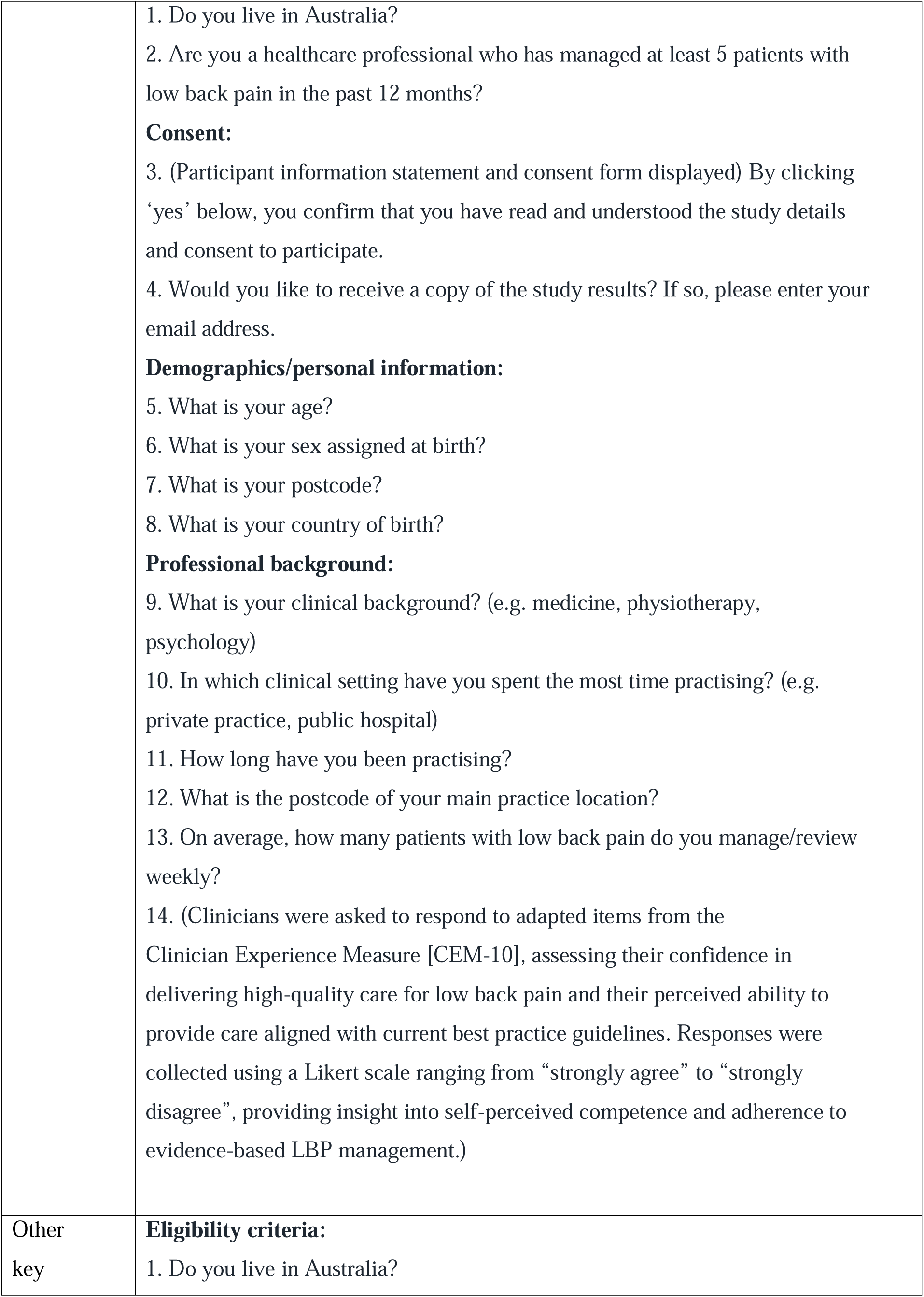
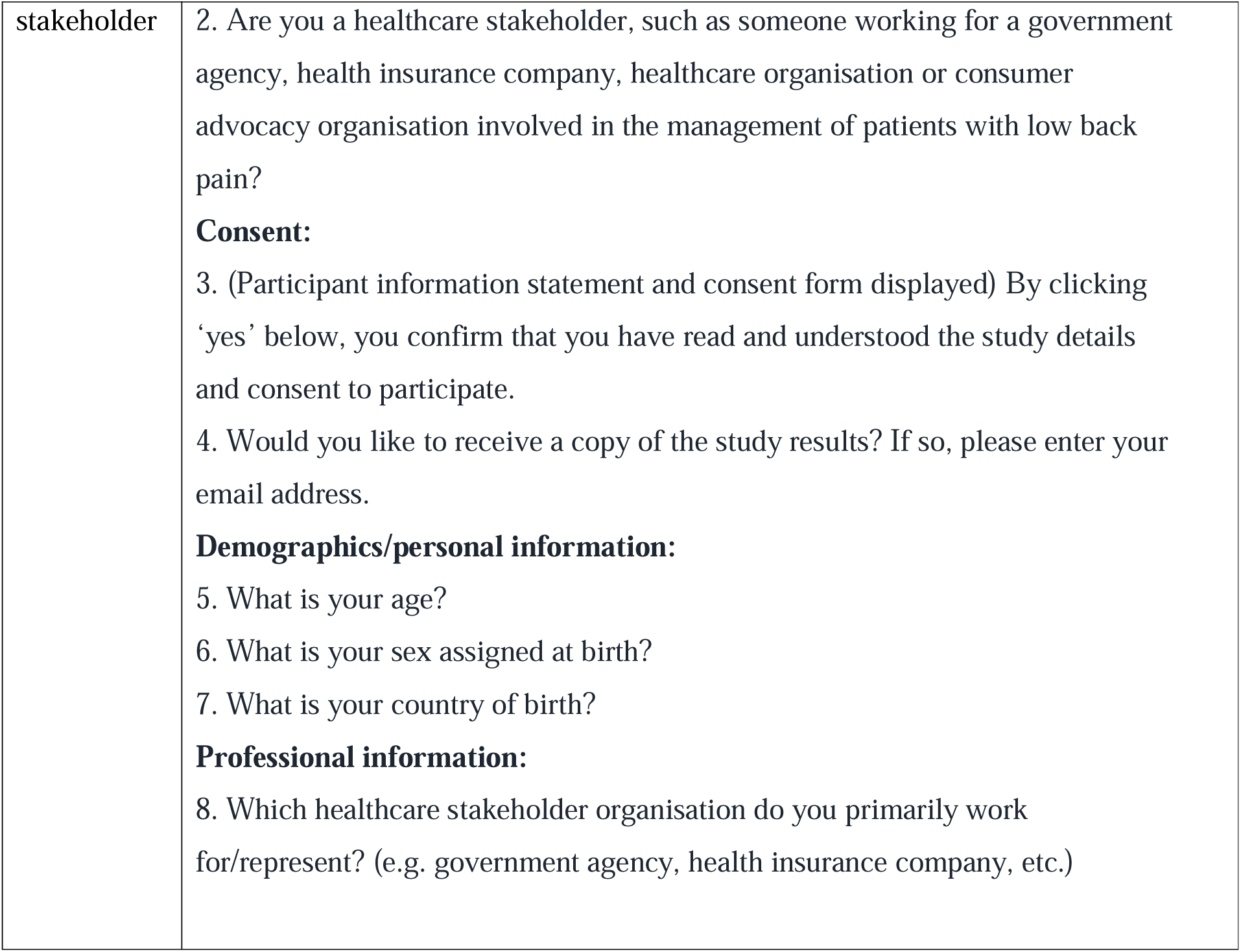
Summary of pre-interview survey questions by participant group.

**Appendix 3:** Summary of interview guides by participant group.

| **Appendix 3: Summary of interview guides by participant group** |  |
| --- | --- |
| **Group** | **Interview guide sections (bolded) and questions (bulleted)** |
| Patient | **Experiences with low back pain:**<br>☐ Can you describe your experience with low back pain?<br>○ When did you first experience it?<br>○ How does it affect your daily life and activities?<br>☐ What treatments have you tried for low back pain?<br>○ Have you tried physiotherapy or other non-surgical options before being referred?<br>○ How effective did you find these treatments?<br>☐ What challenges have you faced in managing your low back pain?<br>○ What would have made it easier to manage your low back pain or access care?<br>**Experiences with referral to spine surgery clinics:**<br>☐ Can you walk me through the process of being referred to a spine surgery clinic?<br>○ Who referred you?<br>○ What was your understanding of why you were referred?<br>☐ How did you feel about the referral process?<br>○ Were any parts confusing, stressful, or unclear?<br>○ Was it what you expected?<br>☐ What information did you receive about the referral, and what to expect from the spine surgery clinic?<br>○ Did you feel adequately informed about what would happen next?<br>**Perceptions and acceptability of the virtual physiotherapist-led triage and management service:**<br>☐ What do you think about the idea of having physiotherapy assessment and treatment via Zoom before seeing a spine surgeon?<br>○ Do you think this is a good way to triage patients, or would you prefer to see a surgeon directly first, or a physiotherapist in-person first? What makes you feel that way? |
|  | ☐ How would you feel about the idea of receiving physiotherapy treatment while you are waiting to see a surgeon?<br>○ Does this approach seem helpful or potentially frustrating?<br>○ Would knowing that you will have physiotherapy in the meantime change how you feel about waiting for surgery?<br>○ Do you think receiving physiotherapy during the waiting period could help improve your condition, mental health or possibly prevent you from having surgery?<br>☐ What are your thoughts on the use of Zoom for physiotherapy treatment, especially if you are on a waiting list to see a spine surgeon?<br>○ Do you feel comfortable using Zoom for your consultation?<br>○ Do you think virtual physiotherapy is as effective as in-person physiotherapy?<br>☐ Would you feel confident in the recommendations made by a physiotherapist regarding whether you should continue with physiotherapy or see a surgeon, even if you haven't yet seen a surgeon? Why or why not?<br>○ Do you trust that the physiotherapist can make the right decision about your treatment without the surgeon's input? Does this cause any fear or anxiety?<br>☐ Do you think a collaborative approach between physiotherapists and spine surgeons is a better way to make treatment decisions?<br>○ Does knowing that both the physiotherapist and surgeon are involved in decision-making make you feel more confident in the process?<br>☐ The idea and design of our proposed model of care came from consultation with patients who have low back pain and clinicians who treat them. Would this affect your opinion about its value and likely effectiveness?<br>☐ What improvements, if any, would you suggest for this proposed model of |
|  | care to make it more acceptable, effective and efficient for people with low back pain?<br>○ Are there any aspects of the process that you think could be streamlined, made more accessible, or improved in terms of support? |
| Clinician | **Experiences with low back pain management:**<br>☑ Can you describe your experience in managing patients with low back pain?<br>○ What are some of the challenges you commonly face in treating this condition?<br>☑ What treatments or interventions do you commonly recommend to patients with low back pain?<br>○ How effective have you found these treatments to be in your practice?<br>☑ What role, if any, does collaboration with other clinicians play in your management of people with low back pain?<br>**Experiences with referrals to spine surgery clinics:**<br>☑ Can you describe your experience with referring patients to spine surgery clinics?<br>○ What factors influence your decision to refer a patient for surgery?<br>☑ How do you feel about the referral pathway to spine surgery clinics for low back pain patients?<br>○ Are there any aspects of the process that you find confusing, frustrating or unclear?<br>☑ What information do you typically provide to patients when referring them to a spine surgery clinic?<br>○ Do you think patients are generally well-informed about the next steps and the purpose of the referral?<br>**Perceptions and acceptability of the virtual physiotherapist-led triage and** |
|  | **management service:**<br>☐ How do you think physiotherapists' involvement in triaging and managing patients before they see a spine surgeon could impact the healthcare system?<br>○ What do you think are the main benefits or risks of having physiotherapists leading the triage and management process for low back pain patients?<br>☐ In your opinion, how effective is telehealth for physiotherapy consultations compared to in-person physiotherapy?<br>○ Are there particular challenges or benefits you see with telehealth physiotherapy for low back pain?<br>☐ What improvements, if any, would you suggest to enhance the implementation or adoption of the proposed new model of care?<br>○ In your opinion, what could improve the effectiveness or efficiency of the service?<br>☐ In your opinion, how could this service be integrated into current clinical workflows and improve patient care?<br>○ What do you think would be the main challenges or barriers in integrating this new model of care into existing clinical workflows?<br>○ What would you propose to address these challenges or barriers?<br>○ What specific skills or training would be most important for physiotherapists to effectively triage patients for surgical or non-surgical care? |
| Other key stakeholder | **Organisational experience with low back pain management:**<br>☐ Can you describe your organisation's role in the management of patients with low back pain?<br>○ What are the main services, policies or initiatives your organisation offers that impact people with low back pain?<br>○ Does your organisation collaborate with other stakeholders in the management of people with low back pain? |
|  | ☐ What challenges does your organisation face in the management of people with low back pain at the healthcare system or population level?<br>☐ What kind of information does your organisation rely on to make decisions about the management of people with low back pain?<br>○ How do you use this information to guide institutions' policies and/or intervention strategies?<br>**Perspectives on the referral process to spine surgery clinics:**<br>☐ How do you feel about the current referral process to spine surgery clinics for low back pain patients?<br>○ Are there any aspects of the referral process that you think need improvement?<br>○ Does your organisation have any policies, recommendations or guidelines about when and how people should be referred to a spine surgery clinic?<br>☐ How does your organisation view the role of physiotherapy and non-surgical treatments in managing people with low back pain prior to referral to surgery?<br>○ Do you have any policies, recommendations or initiatives in place to promote physiotherapy as a first-line assessment and management of patients with low back pain?<br>**Perceptions and acceptability of the virtual physiotherapist-led triage and management service:**<br>☐ What do you think about the idea of having a physiotherapist leading the triage process for low back pain patients via telehealth before they see a spine surgeon?<br>○ Do you have any concerns about this proposed model of care?<br>○ Would you support or promote this model of care within your organisation?<br>○ Do you believe telehealth physiotherapy can be as effective as in-person physiotherapy for managing people with low back pain? |
|  | Why or why not?<br>☐ What impact do you think the introduction of a virtual physiotherapist-led triage service could have on the broader healthcare system?<br>○ Do you see it as a potential solution for reducing wait times, improving access to care, or enhancing the overall patient experience? Why or why not?<br>○ How might your organisation's policies align with or support this new model of care?<br>☐ How do you think the integration of a physiotherapist-led triage process might affect the coordination of care and communication between healthcare providers?<br>○ Do you think this collaborative model between physiotherapists and surgeons could improve the decision-making process for patient management?<br>☐ Are there any potential barriers or challenges that you see for health services in implementing this virtual physiotherapist-led triage model?<br>☐ What improvements, if any, would you suggest for this new service to make it more effective or easier to implement at the organisational level?<br>○ Are there any adjustments that would make it more feasible or acceptable to stakeholders, including patients, healthcare providers or insurance companies (for example)? |

